# Cardiopulmonary Exercise Tests in People with Chronic Stroke: Interpretation and Clinical Application

**DOI:** 10.1101/2024.10.16.24315527

**Authors:** Kiersten M. McCartney, David Edwards, Ryan Pohlig, Pierce Boyne, Tamara Wright, Henry Wright, Matthew Overstreet, Darcy S. Reisman

## Abstract

**Objective:** To understand in people with stroke: 1) reasons for cardiopulmonary exercise test termination, 2) how frequently secondary criteria indicating a maximal test are met, and 3) how additional clinical measures provide context to interpreting test data.

**Design:** A secondary analysis from the Promoting Recovery Optimization of Walking Activity in Stroke (NCT02835313) clinical trial.

**Setting:** Four outpatient rehabilitation clinics.

**Participants:** People with chronic stroke able to walk without assistance of another person.

**Intervention:** Participants (*n* = 250) were randomized into a high-intensity treadmill or a step activity monitoring group. Cardiopulmonary exercise tests were conducted pre- (*n* = 247) and post-intervention (*n* = 185).

**Main Measures:** The primary measure was reason for cardiopulmonary exercise test termination. Secondary measures included: oxygen consumption, ventilatory threshold, peak heart rate, respiratory exchange ratio, six-minute walk test, and fastest walking speed. General Linear Mixed Methods were used to determine the fixed main effects of group and time, and their interaction for outcome variables.

**Results:** There were 4 categories of test termination. Only 1.9% of tests achieved the threshold to confirm a maximal aerobic effort. Despite one group receiving a high-intensity walking intervention, there were no between group differences in oxygen consumption or ventilatory threshold. There were significant differences between groups in measures of walking capacity.

**Conclusions:** Analyses indicate few with chronic stroke demonstrate a maximal aerobic effort on a cardiopulmonary exercise test. If the cardiorespiratory system is not thoroughly taxed during these tests in people with chronic stroke, interpreting results as their cardiorespiratory fitness should be done cautiously.

**Clinical Messages:**

- People with stroke rarely (< 2%) attain an aerobic maximum on cardiopulmonary exercise tests.
- People with chronic stroke, frequently stop these tests due to biomechanical reasons which could reflect neuromuscular impairments, not cardiorespiratory impairments.
- Interpretation of test results as a measure of solely cardiorespiratory fitness or to prescribe exercise should be done cautiously in people with chronic stroke.

## INTRODUCTION

Maximum oxygen consumption is a measure of the body’s maximum capacity to utilize oxygen during incremental exercise and is typically interpreted as a measure of an individual’s cardiorespiratory fitness. Cardiopulmonary exercise tests, with measurements of oxygen and carbon dioxide concentration, are considered the gold standard to quantify cardiorespiratory fitness.^1, 2^

The American College of Sports Medicine has set criteria to guide termination of a cardiopulmonary exercise test, including absolute and relative indications and participant desire to stop or inability to continue the test.^3^ Additionally, there are several defined secondary criteria to confirm a maximum aerobic effort, and thus maximum oxygen consumption, was achieved during testing.^3^ These criteria can be used in combination to confirm maximum oxygen consumption was attained during the test.^3, 4^

Prior literature demonstrates people with chronic stroke have peak oxygen consumption values which are 53% of those found in age- and sex-matched healthy controls.^5^ This appears to indicate people post-stroke have significantly reduced cardiorespiratory fitness. However, people with chronic stroke have multiple impairments across various physiological systems including reduced neuromotor function which significantly affects walking ability^5^. This suggests that there are a variety of reasons why a cardiopulmonary exercise test may be terminated in this population *prior to* reaching a *maximum <u>aerobic</u> effort*. If cardiopulmonary exercise tests in people with chronic stroke are stopped for reasons other than previously described criteria, the impact of this on resultant data needs to be understood for appropriate clinical application.

Previous literature has provided limited information on why cardiopulmonary exercise tests are terminated in those post-stroke, and if secondary criteria to deem a maximal aerobic effort were reached.^6, 7^ It is recommended individuals with stroke complete a cardiopulmonary exercise test prior to engaging in aerobic exercise program, therefore understanding the results from these tests is imperative to apply to clinical practice.^8–10^

The purpose of this study was to 1) quantify the reason and frequency of termination categories for cardiopulmonary exercise tests, 2) quantify how frequently secondary criteria for a maximal aerobic effort are met, and 3) determine how additional cardiorespiratory measures in combination with clinical provide context to interpreting cardiopulmonary exercise test results in people with chronic stroke following an aerobic exercise intervention.

## METHODS

### Study Design and Participants

This secondary analysis leveraged data from the large (*n* = 250) multi-site, randomized clinical trial “*Promoting Recovery Optimization with WALKing Exercise after Stroke*” (NCT02835313). The “*Promoting Recovery Optimization with WALKing Exercise after Stroke*” full methods, protocol and main analyses have previously been reported.^11, 12^ The information provided here is limited to that which is necessary to address the purpose of this analysis. To be included in the “*Promoting Recovery Optimization with WALKing Exercise after Stroke*” study, participants demonstrated: (1) Age between 21-85 years, (2) ≥ 6-months post-stroke, (3) ability to walk at self-selected speed without assistance from a person, (4) self-selected walking speed 0.3-1.0 meters per second (5) resting heart rate between 40-100 beats per minute, and (6) resting blood pressure between 90/60 to 170/90.^11, 12^ Participants were excluded from the study if they demonstrated: (1) evidence of cerebellar stroke, (2) secondary neurologic conditions, (3) lower limb Botulinum toxin injection in prior 4 months, (4) current participation in physical therapy, (5) inability to walk outside the home prior to stroke, (6) coronary artery bypass graft, stent placement or myocardial infarction within past 3 months, (7) musculoskeletal pain which limits activity, (8) inability to communicate with investigators, and/or (9) score >1 on question 1b and >0 on question 1c on the National Institutes of Health Stroke Scale.^11, 12^

### Baseline Examinations

During a baseline evaluation participants completed clinical measurements, treadmill acclimatization, and a symptom limited, maximal treadmill cardiopulmonary exercise test with concurrent, continuous electrocardiogram monitoring. Demographic and medical information including age, sex, use of beta-blocker medication, and time since initial stroke was collected. Self-selected walking speed and fastest walking speed were captured through the 10-meter walk test, and walking endurance through the six-minute walk test. The 10-meter walk test and six-minute walk test are valid and reliable measures of walking speed and walking endurance, respectively, in people with chronic stroke.^8, 13–16^ The six-minute walk test is a sub-maximal test which quantifies an individual’s capacity to walk for extended periods of time,^16^ and is the strongest measure to distinguish between individuals with stroke who are home versus community ambulators. Given that there are a variety of reasons why a cardiopulmonary exercise test may be terminated in someone with stroke, it is important to examine data from the cardiopulmonary exercise test in conjunction with other clinical measures of walking capacity for a more comprehensive understanding of cardiopulmonary exercise test data and its application to clinical practice.

Treadmill acclimatization took place following the over-ground walking tests to familiarize subjects and determine treadmill speed for the cardiopulmonary exercise test. An overhead harness was used for safety and did not provide any off-loading of body weight. Participants were allowed to use handrails and normal orthotic devices. Treadmill speed began at 0.2 miles per hour below the individual’s average overground self-selected walking speed. Every 10-15 seconds, treadmill speed was increased by 0.1 miles per hour until: subjects were unable to continue at current pace due to biomechanical faults (such as toe-catching, drifting back on the treadmill with inability to correct forward, or a significant increase in handrail support to support an upright position), and/or if subject’s heart rate exceeded the following:

> (((220-age) – Resting heart rate) × 80%) + Resting heart rate.

The cardiopulmonary exercise test treadmill speed was set at 85% of the maximal treadmill acclimatization speed.

### Cardiopulmonary Exercise Testing

If eligible, participants were scheduled for a symptom limited, maximal treadmill cardiopulmonary exercise test under the supervision of a Nurse Practitioner, Exercise Physiologist(s), and Physical Therapist. Participants body mass index was recorded, and a 12- lead electrocardiogram was placed. After participants remained in a seated, resting position for a minimum of 5 minutes, resting vital signs were measured. Participants were then instructed to stand, and vital sign measurement was repeated. Standard instructions were provided for the cardiopulmonary exercise test, emphasizing this test was intended to be a maximal effort with participants instructed to walk until they were unable to continue walking or to let study personnel know if they needed to stop the test for any other reason.

Participants were directed to the treadmill and sat on a chair to be fit with an overhead safety harness and a non-rebreathing, airtight facemask over the nose and mouth. Upon proper facemask fitting, respiratory measurements (TrueOne 2400 (Parvo medics, Salt Lake City, Utah); K4b2 Portable Metabolic System (COSMED, Rome, Italy)) began recording continuously, averaging measurements into 15-second intervals. Resting measurements were obtained for at least the first 3-5 minutes until oxygen consumption stabilized. Participants were instructed to stand, and the safety harness was adjusted to ensure no body weight support. The treadmill started at 0.1 miles per hour and within 45 seconds, the speed increased to 85% of the individual’s maximal treadmill acclimatization speed. Treadmill speed remained constant for the remainder of the test. Participants walked at 0% incline for 2 minutes, the incline increased to 2% for the following 2 minutes, and each minute thereafter the incline was increased by 2% until test termination. Heart rate and electrocardiogram were monitored throughout with blood pressure taken during the last 15 seconds of each test phase. The cardiopulmonary exercise test was terminated if: participants requested to stop, observable biomechanical faults prevented participants from walking safely, and/or if other absolute termination criteria, per American College of Sports Medicine guidelines, were reached.^17^ Following termination, the treadmill was stopped and one minute of recovery cardiorespiratory measurements were recorded in a resting position prior to facemask removal. Blood pressure, heart rate, and electrocardiogram continued to be monitored until the Nurse Practitioner deemed it was safe to stop. The reason for the cardiopulmonary exercise test termination was recorded (Stop Category) and all cardiopulmonary exercise test results were reviewed by the study cardiologist.

Following American College of Sports Medicine recommendations, participants were deemed to have provided a maximal aerobic effort on the cardiopulmonary exercise test if they reached an respiratory exchange ratio value of greater than or equal to 1.10 and were within 10 beats per minute of their age-predicted heart rate maximum.^2^ Age-predicted heart rate max was calculated as:

> 208 – (0.7 x age)^18^

with participants maximal percentage of age-predicted heart rate max calculated as:

> (Heart Rate _max_ / age-predicted heart rate maximum) x 100%.

As there is controversy on the appropriate thresholds for these secondary criteria in aging and/or clinical populations, several thresholds for heart rate and respiratory exchange ratio were evaluated in this population. Blood lactate measurements and rate of perceived exertion were not analyzed as secondary criteria as these measurements were not recorded. *Ventilatory Threshold Determination*

Ventilatory threshold is considered the intensity at which ventilation begins increasing at a rate faster than oxygen is consumed.^19^ It has been theorized ventilatory threshold may more accurately represent cardiorespiratory fitness and the ability to withstand continuous submaximal work,^20^ including in people with chronic stroke.^21^ Ventilatory threshold was automatically and manually determined using the V-Slope and Excess carbon dioxide methods.^22, 23^ The automated and manual oxygen consumption (mL/kg/min) values at ventilatory threshold for each method were independently averaged to provide a final manual value of oxygen consumption at ventilatory threshold and automated value of oxygen consumption at ventilatory threshold. The *mcp* package in R was used for automatic determination and three independent investigators (KM, DE, MO) visually determined ventilatory threshold from 50 randomly selected cardiopulmonary exercise tests for reliability analyses. The automated oxygen consumption at ventilatory threshold values were used for analyses (*see Appendix*).

### Interventions

The protocol design has been published.^11, 12^ Briefly, participants were enrolled and randomized to one of three intervention groups: high-intensity treadmill walking, step-activity monitoring, or a high-intensity treadmill walking and step-activity monitoring combined intervention.^12^ All interventions consisted of 2-3 training sessions/week for 12 weeks. The high-intensity treadmill walking intervention utilized treadmill walking with a goal of accumulating as many minutes as possible (maximum of 30 minutes/session) at or above 70% heart rate reserve. The step-activity monitoring intervention utilized step monitoring with a Fitbit and individualized goal setting to progressively increase participants’ average daily step activity. The high-intensity treadmill walking and step-activity monitoring group received both interventions across the intervention period.

### Statistical Analysis

All statistical analyses were conducted using Statistical Package for the Social Sciences (SPSS) (Version 29.0; IBM, Armonk, New York). Cardiorespiratory data was processed by a research team member at UD (KM). Respiratory exchange ratio and oxygen consumption values at rest and max, were defined as the average two time points (30 seconds total) prior to standing or cardiopulmonary exercise test termination, respectively. Participant characteristics and baseline measurements of walking speed and endurance were compared between treatment intervention groups using one-way analysis of variance for continuous variables and Fisher exact tests for categorical variables. Descriptive analyses were used for Stop Category and secondary criteria which confirm a maximum aerobic effort. General Linear Mixed Methods were used to determine the fixed, main effects of group and time, and their interaction for resting oxygen consumption, oxygen consumption at ventilatory threshold, maximum oxygen consumption, and length of cardiopulmonary exercise test.

## RESULTS

### Participants

Participants (*n* = 250) were randomized into 1 of 3 intervention groups: (1) high-intensity treadmill walking (*n* = 89), (2) step-activity monitoring (*n* = 81), or (3) high-intensity treadmill walking and step-activity monitoring (*n* = 80). A total of 432 cardiopulmonary exercise tests were conducted across groups, with 247 cardiopulmonary exercise tests conducted at baseline and 185 cardiopulmonary exercise tests at post. As there were no baseline differences between the two groups which received the high-intensity treadmill exercise intervention (high-intensity treadmill walking, high-intensity treadmill walking and step-activity monitoring), these groups were pooled (referred to as “HI-Tread” for remainder of text) and compared to the group that did **not** receive a high-intensity exercise intervention (step-activity monitoring; “NO-Tread”). The only baseline between group difference was a higher body mass index in the NO-Tread group (*p* = 0.04). The General Linear Mixed Methods with robust errors controlled for this difference in BMI. Descriptive characteristics are in Table 1.

**Table 1.** Participant Characteristics.

| **Participant Characteristics** |  |  |
| --- | --- | --- |
|  | **HI-Tread (*n* = 169)** | **NO-Tread (*n* = 81)** |
| Age (years) | 63.1 ± 12.5 | 62.2 ± 12.9 |
| Sex, *n* (% Female) | 77 (45.6) | 39 (48.1) |
| Side of Hemiparesis (*n*, %) |  |  |
| Left | 82 (59.8) | 42 (51.9) |
| Right | 80 (47.3) | 34 (47.3) |
| Bilateral | 7 (4.1) | 5 (4.1) |
| Time Since Initial Stroke (months) | 44.5 ± 56.1 | 46.8 ± 73.2 |
| Body mass index (kg/m<sup>2</sup>) | 29.8 ± 6.4** | 31.6 ± 7.0** |
| Beta-Blocker Medication, *n* (% Yes) | 68 (40.2) | 35 (43.2) |
| Self-selected walking speed (m/s) | 0.70 ± 0.2 | 0.68 ± 0.2 |
| Fast walking speed (m/s) | 1.00 ± 0.3 | 0.95 ± 0.3 |
| 6-Minute Walk Test (m) | 289.7 ± 117.3 | 279.2 ± 110.1 |
| Average Steps/Day | 3869.4 ± 2042.0 | 3844.8 ± 2192.6 |
| Assistive Device Use, *n* (% Yes) | 82 (48.5) | 47 (58.0) |
| Orthotic Device Use, *n* (% Yes) | 43 (25.4) | 25 (30.9) |
| Ethnicity, *n* (% Hispanic or Latino) | 8 (4.7) | 2 (2.5) |
| Race (*n*, % sample) |  |  |
| Native American/American Indian/Alaska Native | 7 (4.1) | 4 (4.9) |
| Asian | 12 (7.1) | 2 (2.5) |
| Native Hawaiian or Pacific Islander | 0 (0) | 0 (0) |
| African American/Black | 45 (26.6) | 27 (33.3) |
| Caucasian/White | 112 (66.3) | 52 (64.2) |
| Prefer to not Respond | 2 (1.2) | 0 (0) |

*Continuous data presented as mean ± standard deviation.*
*Abbreviations: HI-Tread = high intensity treadmill exercise group, NO-Tread = no treadmill*
*exercise group*
*\*\*significant difference ( $p < 0.05$ )*

### Cardiopulmonary Exercise Test Termination Categories

Cardiopulmonary exercise tests were terminated for 4 main reasons: (1) electrocardiogram abnormalities; (2) abnormal blood pressure or heart rate response; (3) biomechanical faults that prevented safe continuation of test, such as an inability to keep up with treadmill speed or toe catching; or (4) the participant self-selected to terminate the test. Examples of reasons participants elected to terminate the test included: discomfort with the mask, a lack of desire to continue walking, full body fatigue, excessive upper or lower extremity fatigue on the hemiparetic side, and fear or anxiety. Only two cardiopulmonary exercise tests were terminated for reasons outside of these 4 categories, with both tests stopped due to excessive shortness of breath. There were no significant differences between HI-Tread and NO-Tread at baseline or post on reason for cardiopulmonary exercise test termination (*p* > 0.146). Collectively across both timepoints, cardiopulmonary exercise tests were terminated 14.7% due to abnormal electrocardiograms, 17.7% due to abnormal blood pressure or heart rate, 24.9% due to biomechanical faults, and 43.0% due to Self-Selected stop (Table 2).

**Table 2.** Cardiopulmonary Exercise Test Stop Categories.

| **Group** | **CPET Stop Categories** |  |  |
| --- | --- | --- | --- |
|  | **HI-Tread** | **NO-Tread** | **ALL** |
| **ECG** (*n, %*) |  |  |  |
| Baseline | 26 (15.6) | 14 (17.5) | 40 (16.2) |
| Post | 15 (12.2) | 8 (12.9) | 23 (12.4) |
| **BP/HR** (*n, %*) |  |  |  |
| Baseline | 25 (15.0) | 13 (16.3) | 38 (15.4) |
| Post | 30 (24.4) | 8 (12.9) | 38 (20.5) |
| **Biomechanical** (*n, %*) |  |  |  |
| Baseline | 47 (28.1) | 23 (28.7) | 70 (28.3) |
| Post | 26 (21.1) | 11 (17.7) | 37 (20.0) |
| **Self-Selected** (*n, %*) |  |  |  |
| Baseline | 68 (40.7) | 30 (37.5) | 98 (39.7) |
| Post | 53 (43.1) | 34 (54.8) | 87 (47.0) |

*Abbreviations: HI-Tread = high intensity treadmill exercise group; NO-Tread = no treadmill*
*exercise group; ECG = electrocardiogram; BP = blood pressure; HR = heart rate*

### Cardiopulmonary Exercise Test Data

There were no significant main effects of group or time for resting oxygen consumption (*p*=0.623, *p*=0.491), ventilatory threshold (*p*=0.051, *p*=0.682), or maximal oxygen consumption (*p*=0.076, *p*=0.498) between those who underwent a high-intensity treadmill walking exercise intervention (HI-Tread) and those who did not (NO-Tread) (Table 3). However, there was a significant difference between groups in the change of cardiopulmonary exercise test length over time (*p* < 0.001). The HI-Tread group’s cardiopulmonary exercise test CPET length improved by 1.92 minutes (95% CI = 1.493-2.353, *p* < 0.001), while the NO-Tread group’s cardiopulmonary exercise test length did not change (0.38 minutes; *p* = .107).

**Table 3.** Frequency Secondary Criteria Thresholds Met.

| **Secondary Criteria Met for Maximal Aerobic Effort** |  |  |  |
| --- | --- | --- | --- |
| **Group** | **HI-Tread** | **NO-Tread** | **All** |
| **Peak HR:** |  |  |  |
| **Within 10bpm APHRM (n, %)** |  |  |  |
| *Baseline* | 13 (7.7) | 2 (2.5) | 15 (6.1) |
| *Post* | 15 (8.9) | 4 (4.9) | 19 (10.3) |
| **≥ 95% APHRM (n, %)** |  |  |  |
| *Baseline* | 13 (7.7) | 1 (1.2) | 14 (5.7) |
| *Post* | 14 (11.1) | 4 (6.3) | 18 (9.7) |
| **≥ 90% APHRM (n, %)** |  |  |  |
| *Baseline* | 23 (13.6) | 6 (7.4) | 29 (11.7) |
| *Post* | 24 (19.0) | 5 (7.9) | 29 (15.7) |
| **Peak RER:** |  |  |  |
| **≥ 1.1 (n, %)** |  |  |  |
| *Baseline* | 35 (20.7) | 11 (13.6) | 46 (18.6) |
| *Post* | 24 (19.5) | 10 (16.1) | 34 (18.4) |
| **≥ 1.0 (n, %)** |  |  |  |
| *Baseline* | 76 (45.5) | 34 (42.5) | 110 (44.5) |
| *Post* | 64 (52.0) | 30 (48.4) | 94 (50.8) |
| **Both Peak HR within 10bpm of APHRM and RER ≥ 1.1 (n, %)** |  |  |  |
| *Baseline* | 3 (1.8) | 0 (0) | 3 (1.2) |
| *Post* | 4 (2.4) | 1 (1.2) | 5 (2.7) |
| **Both ≥ 90% APHRM and RER ≥ 1.0 (n, %)** |  |  |  |
| *Baseline* | 11 (6.6) | 3 (3.8) | 14 (5.7) |
| *Post* | 18 (14.6) | 2 (3.2) | 20 (10.8) |

*Data presented as n (%). Abbreviations: HR = heart rate; CPET = cardiopulmonary exercise test; APHRM = age-predicted heart rate maximum, calculated as $208 - (0.7 \times \text{age})$ ; RER = respiratory exchange ratio*

### Secondary Criteria

Of the cardiopulmonary exercise tests conducted at baseline, 15 participants (6.1%) reached a peak heart rate within 10 beats per minute (bpm) of age-predicted heart rate maximum and 46 participants (18.6%) reached an respiratory exchange ratio of ≥ 1.1. At post, these percentages were similar as19 participants (10.3%) reached a peak heart rate within 10 bpm of age-predicted heart rate maximum and 34 participants (18.4%) reached an respiratory exchange ratio of ≥ 1.1. The threshold of **both** secondary criteria was met in only 8 (1.9%) of the 432 cardiopulmonary exercise tests conducted (Table 4). When less strict threshold criteria (a heart rate of ≥ 90% age-predicted heart rate maximum and an respiratory exchange ratio ≥ 1.0) were used, only 7.9% of all tests met the criteria. Due to the very limited number of tests that could be quantified as a maximal aerobic effort, the highest oxygen consumption value obtained in these cardiopulmonary exercise tests cannot be accepted as a maximal oxygen consumption, and thus, will be referred to as a peak oxygen consumption for the remainder of this analysis.

**Table 4.** Cardiopulmonary Exercise Test Data.

| **CPET Data** |  |  |
| --- | --- | --- |
|  | **HI-Tread** | **NO-Tread** |
| **Rest VO<sub>2</sub> (mL/kg/min)** |  |  |
| *Baseline* | 2.7 ± 0.9 | 2.7 ± 0.9 |
| *Post* | 2.8 ± 0.9 | 2.5 ± 0.8 |
| **Peak VO<sub>2</sub> (mL/kg/min)** |  |  |
| *Baseline* | 12.8 ± 5.0 | 12.1 ± 4.2 |
| *Post* | 13.3 ± 5.8 | 12.1 ± 4.2 |
| **VT-VO<sub>2</sub> (mL/kg/min)** |  |  |
| *Baseline* | 9.4 ± 3.2 | 8.8 ± 2.7 |
| *Post* | 9.5 ± 3.6 | 8.7 ± 2.9 |
| **% of peak VO<sub>2</sub>** |  |  |
| *Baseline* | 76.7 ± 15.5 | 74.9 ± 12.5 |
| *Post* | 74.7 ± 18.0 | 73.2 ± 13.4 |
| **Length of Test (min)** |  |  |
| *Baseline* | 6.0 ± 2.8 | 5.6 ± 2.7 |
| *Post* | 8.0 ± 3.3** | 6.0 ± 2.6** |
| **Peak Heart Rate (bpm)** |  |  |
| *Baseline* | 126.0 ± 21.6 | 123.0 ± 20.8 |
| *Post* | 126.4 ± 23.0 | 123.1 ± 21.9 |
| **% of APhRM** |  |  |
| *Baseline* | 77.3 ± 12.3 | 75.2 ± 11.9 |
| *Post* | 77.8 ± 13.0 | 75.5 ± 12.4 |
| **Respiratory Exchange Ratio** |  |  |
| *Baseline* | 0.99 ± 0.13 | 0.97 ± 0.12 |
| *Post* | 1.00 ± 0.13 | 0.99 ± 0.12 |

Data presented as mean ± SD. Abbreviations: CPET = cardiopulmonary exercise test; VO<sub>2</sub> = volume of oxygen consumption; VT = ventilatory threshold; APhRM = age-predicted heart rate maximum, calculated as 208 – (0.7 \* age); RER = respiratory exchange ratio
\*\* = significant ( $p < 0.001$ ) difference between groups

### Clinical Walking Tests

There was a significant difference between groups in the change of six-minute walk test distance over time (*p* = 0.045). While both groups improved over time, the HI-Tread group’s six-minute walk test distance improved by 41.59 meters (95% Confidence Interval = 31.851-51.327, *p* < 0.001), while the NO-Tread group’s six-minute walk test distance improved by 25.94 meters (95% Confidence Interval = 14.126-37.754, *p* < 0.001). Additionally, there was a significant difference between groups in the change of fastest walking speed over time (*p* = 0.028), where the HI-Tread group’s fastest walking speed improved by 0.17 meters per second (95% Confidence Interval = 0.142-0.202, *p* < 0.001) and the NO-Tread group’s fastest walking speed improved by 0.12 meters per second (95% Confidence Interval = 0.085-0.156, *p* < 0.001). For self-selected walking speed, both groups improved over time (*p* < 0.001) with no difference between groups (*p* < 0.293). The HI-Tread’s self-selected walking speed improved by 0.14 meters per second (95% Confidence Interval = 0.085-0.147, *p* < 0.001) and NO-Tread’s by 0.12 meters per second (95% Confidence Interval = 0.113-0.159, *p* < 0.001).

## DISCUSSION

Cardiopulmonary exercise tests are designed to test how multiple systems interact to deliver and utilize oxygen through an incrementally increased workload and are considered the gold standard to assess a person’s cardiorespiratory fitness. In recent years, there has been an increase in the number of studies using cardiopulmonary exercise tests to quantify cardiorespiratory fitness in people with chronic stroke.^5, 6, 24–31^ This secondary analysis sought to provide a better understanding of how to interpret cardiopulmonary exercise test data in people with chronic stroke by analyzing measures taken directly from the cardiopulmonary exercise test in conjunction with clinical waking capacity tests. These analyses provide a more comprehensive understanding of what impairments may be impacting performance on cardiopulmonary exercise tests in people with chronic stroke.

The results of this study revealed that over 50% of cardiopulmonary exercise tests in those with chronic stroke terminate for reasons other than participant’s self-selecting to end the test. Over 20% of tests were stopped due to biomechanical faults, such as an inability to maintain speed on the treadmill or toe catching. Additionally, less than 2% of all tests met secondary criteria to deem a maximal aerobic effort was provided. There were no differences in meeting secondary criteria between those on beta-blocker medications (42%) versus those who were not on beta-blocker medications (*p* = .342). Recent literature has called into question the thresholds used to deem a maximum aerobic effort, and suggest for these criteria to be adjusted for age and sex.^32^ The lowest respiratory exchange ratio threshold suggested for males and females over age 65 is attaining at least a 1.0. Even by this lower standard, only 8% of the participants reached this threshold. This indicates there is a near complete failure to meet maximal aerobic effort threshold criteria during the cardiopulmonary exercise test. These results strongly suggest the cardiorespiratory system is likely not the primary limiting factor for cardiopulmonary exercise test termination in people with chronic stroke.

Additionally, despite one group (HI-Tread) undergoing a 12-week high intensity walking exercise intervention, there were no significant differences within or between groups over time in ventilatory threshold or peak oxygen consumption. This would indicate a lack of change in cardiorespiratory fitness following an aerobic intervention. It is unlikely this was due to a poor execution of the exercise intervention as the participants in the HI-Tread group walked for 28.4 ± 2.7 (mean ± SD) minutes per session, in 25.2 ± 8.2 sessions across 12-weeks, at an average intensity of 63.6 ± 12.0% heart rate reserve. This intensity falls into the American College of Sports Medicine’s definition of “vigorous” intensity.^3^ Despite a lack of change in ventilatory threshold or peak oxygen consumption, there was a significant difference between groups in the length of cardiopulmonary exercise test. Since treadmill speed on the cardiopulmonary exercise test remained constant between baseline and post evaluations, if the length of the test increases, it indicates the participant can sustain that same speed, plus an increased incline, for a longer duration following the intervention. Despite the lack of change in ventilatory threshold, and no change in the relative time at which ventilatory threshold occurred during the test, people with chronic stroke who received a high-intensity walking intervention were able to achieve a higher absolute workload. This is further supported by the data from the six-minute walk test. Since the duration of the six-minute walk test remains constant, if the distance covered in that time increases, participants must do so by increasing their average walking speed over the course of the test. The significant improvement in the six-minute walk test in this study also demonstrates that the participants were able to maintain a higher overall workload.

If there is a lack of significant change in cardiorespiratory metrics (peak oxygen consumption, ventilatory threshold) following a high-intensity aerobic intervention, this would traditionally indicate a lack of significant change in cardiorespiratory fitness. However, despite a lack of change in cardiorespiratory measurements, there were significant changes in other outcomes (length of cardiopulmonary exercise test, six-minute walk test, fastest walking speed), which indicate that changes to walking capacity did occur. Following a high-intensity aerobic walking intervention, one might expect to see changes in cardiorespiratory metrics, *if the main limiting factor was the aerobic system*. In the present study, individuals with chronic stroke did not have any significant changes in their cardiorespiratory metrics but did have changes in their functional capacity demonstrated by the: ability to walk longer at the same speed (cardiopulmonary exercise test length), walk further in the same amount of time (six-minute walk test), and in their fastest walking speed. This apparent mismatch between cardiorespiratory metrics and other measures may indicate the high-intensity walking intervention broadly improved neuromotor function more than cardiorespiratory function.

Cardiopulmonary exercise tests are designed to test how multiple systems interact to deliver and utilize oxygen through an incrementally increased workload. The data presented here indicate a vast majority of cardiopulmonary exercise tests are terminated prior to reaching a maximal aerobic effort, meaning these values represent a peak oxygen consumption and not a maximal oxygen consumption. It is important then that these results be interpreted as such. Much of the prior literature in people with chronic stroke interprets low peak oxygen consumption values obtained in cardiopulmonary exercise tests as a direct indication of poor cardiorespiratory fitness. However, comprehensively viewing data from the 432 cardiopulmonary exercise tests conducted in this study demonstrate few individuals with chronic stroke meet criteria to deem that a maximal aerobic effort was provided. Thus, the cardiorespiratory system is likely not maximally taxed during cardiopulmonary exercise tests in this population. This suggests the oxygen consumption values obtained from a cardiopulmonary exercise test in people with chronic stroke are not reflective of maximal aerobic capacity, and therefore, cannot be interpreted as just an indication of poor cardiorespiratory fitness.

To our knowledge, this is the largest data set of treadmill cardiopulmonary exercise tests in people with chronic stroke, and with a near equal split of sex (48.1%), which may make our results more representative of this population as compared to previous studies. However, participants had to be able to ambulate without assistance from another person to be included and therefore our results may be an overestimation of peak oxygen consumption values across all people with chronic stroke. The population in this study only included individuals with a self-selected walking speed less than 1.0 meters per second, meaning overall results may not be representative of those with mild or minor motor impairments post-stroke. Previous work has demonstrated poor validity of the rate of perceived exertion scale during high-intensity exercise in people with stroke, therefore the larger clinical trial did not include rate of perceived exertion measurements during the cardiopulmonary exercise test.^33^ As the cardiopulmonary exercise test protocol did not include measurements of rate of perceived exertion and/or lactate, this study is unable to comment on all American College of Sports Medicine secondary criteria.

## CONCLUSIONS

Taken in its totality, the data from the cardiopulmonary exercise tests and from the clinical measures of walking capacity suggest that, in most people with chronic stroke, the cardiorespiratory system may not be the main limiting factor in cardiopulmonary exercise test termination. This directly impacts rehabilitation treatment as a patient’s plan of care is directed at the primary impairments identified through a variety of tests and measures. If the cardiorespiratory system is not thoroughly taxed during a cardiopulmonary exercise test in people with chronic stroke, interpreting results as a representation of an individual’s cardiorespiratory fitness and/or using results to prescribe exercise intensity for an aerobic intervention would likely lead to inappropriate treatment planning.

## Data Availability

All data produced in the present study are available upon reasonable request to the authors.

## References

1. Balady GJ, Arena R, Sietsema K, et al. Clinician’s Guide to cardiopulmonary exercise testing in adults: a scientific statement from the American Heart Association. Circulation 2010; 122: 191–225. 2010/06/30. DOI: 10.1161/CIR.0b013e3181e52e69.

2. Kaminsky LA, Arena R and Myers J. Reference Standards for Cardiorespiratory Fitness Measured With Cardiopulmonary Exercise Testing: Data From the Fitness Registry and the Importance of Exercise National Database. Mayo Clin Proc 2015; 90: 1515–1523. 2015/10/13. DOI: 10.1016/j.mayocp.2015.07.026.

3. Thompson PD, Arena R, Riebe D, et al. ACSM’s new preparticipation health screening recommendations from ACSM’s guidelines for exercise testing and prescription, ninth edition. Curr Sports Med Rep 2013; 12: 215–217. 2013/07/16. DOI: 10.1249/JSR.0b013e31829a68cf.

4. Borg GA. Psychophysical bases of perceived exertion. Med Sci Sports Exerc 1982; 14: 377–381. 1982/01/01.

5. Smith AC, Saunders DH and Mead G. Cardiorespiratory fitness after stroke: a systematic review. Int J Stroke 2012; 7: 499–510. 2012/05/10. DOI: 10.1111/j.1747-4949.2012.00791.x.

6. Woodward JL, Connolly M, Hennessy PW, et al. Cardiopulmonary Responses During Clinical and Laboratory Gait Assessments in People With Chronic Stroke. Phys Ther 2019; 99: 86–97. 2018/11/27. DOI: 10.1093/ptj/pzy128.

7. Duncan PW, Sullivan KJ, Behrman AL, et al. Protocol for the Locomotor Experience Applied Post-stroke (LEAPS) trial: a randomized controlled trial. BMC Neurol 2007; 7: 39. 2007/11/13. DOI: 10.1186/1471-2377-7-39.

8. Winstein CJ, Stein J, Arena R, et al. Guidelines for Adult Stroke Rehabilitation and Recovery: A Guideline for Healthcare Professionals From the American Heart Association/American Stroke Association. Stroke 2016; 47: e98–e169. 20160504. DOI: 10.1161/str.0000000000000098.

9. Billinger SA, Arena R, Bernhardt J, et al. Physical activity and exercise recommendations for stroke survivors: a statement for healthcare professionals from the American Heart Association/American Stroke Association. Stroke 2014; 45: 2532–2553.20140520. DOI: 10.1161/str.0000000000000022.

10. Boyne P, Billinger S, MacKay-Lyons M, et al. Aerobic Exercise Prescription in Stroke Rehabilitation: A Web-Based Survey of US Physical Therapists. J Neurol Phys Ther 2017; 41: 119–128. 2017/03/07. DOI: 10.1097/npt.0000000000000177.

11. Thompson ED, Pohlig RT, McCartney KM, et al. Increasing Activity After Stroke: A Randomized Controlled Trial of High-Intensity Walking and Step Activity Intervention. Stroke 2024; 55: 5–13. 20231222. DOI: 10.1161/strokeaha.123.044596.

12. Wright H, Wright T, Pohlig RT, et al. Protocol for promoting recovery optimization of walking activity in stroke (PROWALKS): a randomized controlled trial. BMC Neurol 2018; 18: 39. 20180412. DOI: 10.1186/s12883-018-1044-1.

13. Sullivan JE, Crowner BE, Kluding PM, et al. Outcome measures for individuals with stroke: process and recommendations from the American Physical Therapy Association neurology section task force. Phys Ther 2013; 93: 1383–1396. 20130523. DOI: 10.2522/ptj.20120492.

14. Eng JJ, Dawson AS and Chu KS. Submaximal exercise in persons with stroke: test-retest reliability and concurrent validity with maximal oxygen consumption. Arch Phys Med Rehabil 2004; 85: 113–118. 2004/02/19. DOI: 10.1016/s0003-9993(03)00436-2.

15. Tyson S and Connell L. The psychometric properties and clinical utility of measures of walking and mobility in neurological conditions: a systematic review. Clin Rehabil 2009; 23: 1018–1033. 20090928. DOI: 10.1177/0269215509339004.

16. Moore JL, Potter K, Blankshain K, et al. A Core Set of Outcome Measures for Adults With Neurologic Conditions Undergoing Rehabilitation: A CLINICAL PRACTICE GUIDELINE. Journal of Neurologic Physical Therapy 2018; 42: 174–220. DOI: 10.1097/npt.0000000000000229.

17. Sietsema KES, William W.; Sue, Darryl Y.; Ward. Wasserman & Whipp’s Principles of Exercise Testing and Interpretation. 6th Ed. ed.: Lippincott Williams & Wilkins (LWW), 2020.

18. Tanaka H, Monahan KD and Seals DR. Age-predicted maximal heart rate revisited. J Am Coll Cardiol 2001; 37: 153–156. 2001/01/12. DOI: 10.1016/s0735-1097(00)01054-8.

19. Franco V. Cardiopulmonary Exercise Testing. In: Vasan RS and Sawyer DB (eds) Encyclopedia of Cardiovascular Research and Medicine. Oxford: Elsevier, 2018, pp.523–526.

20. Anselmi F, Cavigli L, Pagliaro A, et al. The importance of ventilatory thresholds to define aerobic exercise intensity in cardiac patients and healthy subjects. Scandinavian Journal of Medicine & Science in Sports 2021; 31: 1796–1808. DOI: 10.1111/sms.14007.

21. Boyne P, Reisman D, Brian M, et al. Ventilatory threshold may be a more specific measure of aerobic capacity than peak oxygen consumption rate in persons with stroke. Top Stroke Rehabil 2017; 24: 149–157. 20160725. DOI: 10.1080/10749357.2016.1209831.

22. Gaskill SE, Ruby BC, Walker AJ, et al. Validity and reliability of combining three methods to determine ventilatory threshold. Medicine & Science in Sports & Exercise 2001; 33: 1841–1848.

23. Kim KJ, Rivas E, Prejean B, et al. Novel Computerized Method for Automated Determination of Ventilatory Threshold and Respiratory Compensation Point. Front Physiol 2021; 12: 782167. 20211217. DOI: 10.3389/fphys.2021.782167.

24. Billinger SA, Taylor JM and Quaney BM. Cardiopulmonary response to exercise testing in people with chronic stroke: a retrospective study. Stroke Res Treat 2012; 2012: 987637. 2011/10/01. DOI: 10.1155/2012/987637.

25. Dobrovolny CL, Ivey FM, Rogers MA, et al. Reliability of treadmill exercise testing in older patients with chronic hemiparetic stroke. Arch Phys Med Rehabil 2003; 84: 1308–1312. 2003/09/19. DOI: 10.1016/s0003-9993(03)00150-3.

26. MacKay-Lyons MJ and Howlett J. Exercise Capacity and Cardiovascular Adaptations to Aerobic Training Early After Stroke. Topics in Stroke Rehabilitation 2005; 12: 31–44. DOI: 10.1310/RDQM-JTGL-WHAA-XYBW.

27. Globas C, Becker C, Cerny J, et al. Chronic stroke survivors benefit from high-intensity aerobic treadmill exercise: a randomized control trial. Neurorehabil Neural Repair 2012; 26: 85–95. 2011/09/03. DOI: 10.1177/1545968311418675.

28. Mackay-Lyons M. Aerobic treadmill training effectively enhances cardiovascular fitness and gait function for older persons with chronic stroke. J Physiother 2012; 58: 271. 2012/11/28. DOI: 10.1016/s1836-9553(12)70131-5.

29. Munari D, Pedrinolla A, Smania N, et al. High-intensity treadmill training improves gait ability, VO2peak and cost of walking in stroke survivors: preliminary results of a pilot randomized controlled trial. Eur J Phys Rehabil Med 2018; 54: 408–418. 2016/08/31. DOI: 10.23736/s1973-9087.16.04224-6.

30. Boyne P, Dunning K, Carl D, et al. High-Intensity Interval Training and Moderate-Intensity Continuous Training in Ambulatory Chronic Stroke: Feasibility Study. Phys Ther 2016; 96: 1533–1544. 2016/04/23. DOI: 10.2522/ptj.20150277.

31. Holleran CL, Rodriguez KS, Echauz A, et al. Potential Contributions of Training Intensity on Locomotor Performance in Individuals With Chronic Stroke. Journal of Neurologic Physical Therapy 2015; 39: 95–102. DOI: 10.1097/npt.0000000000000077.

32. Edvardsen E, Hem E and Anderssen SA. End criteria for reaching maximal oxygen uptake must be strict and adjusted to sex and age: a cross-sectional study. PLoS One 2014; 9: e85276. 2014/01/24. DOI: 10.1371/journal.pone.0085276.

33. Sage M, Middleton LE, Tang A, et al. Validity of rating of perceived exertion ranges in individuals in the subacute stage of stroke recovery. Top Stroke Rehabil 2013; 20: 519–527. DOI: 10.1310/tsr2006-519.

