## Supplemental Material for "Cardiopulmonary Exercise Tests in People with Chronic Stroke: Interpretation and Clinical Application"

**APPENDIX 1**

**VENTILATORY THRESHOLD METHODOLOGY & RESULTS**

*Ventilatory Threshold Determination*

Ventilatory threshold is considered the point at which ventilation increases at a rate faster than oxygen is consumed. Traditionally, ventilatory threshold is determined through a visual, subjective assessment of expired gas data obtained from cardiopulmonary exercise tests. However, this process is time consuming, requires multiple study personnel to complete, and strict criteria to ensure consistency. Thus, automated analyses have been suggested to provide a consistent and efficient objective analysis of ventilatory threshold with previous literature recommending using the average of multiple ventilatory threshold detection methods to accurately identify ventilatory threshold.^22, 23^ An automated methodology using two common ventilatory threshold determination methods (V-slope and Excess carbon dioxide, respectively) was previously validated in a population of healthy individuals in the United States astronaut corps. This methodology was adopted to test its reliability and validity in this population of people with chronic stroke.^23^

*Automated Ventilatory Threshold Determination*

The V-slope method plots the volume of carbon dioxide production over the volume of oxygen consumed. In this method, ventilatory threshold is defined as the point at which there is an increase in the rate of carbon dioxide production compared to oxygen consumption. In the Excess carbon dioxide method, ventilatory threshold is defined as the time point where there is a sustained rise in carbon dioxide. Using the *mcp* package in R, code was developed to separately determine ventilatory threshold using the V-slope and Excess carbon dioxide methods. The volume of oxygen consumption (mL/kg/min) values at ventilatory threshold for each method were then averaged to provide a final value of volume of oxygen consumption at ventilatory threshold.

*Manual Ventilatory Threshold Determination*

Three independent investigators (KM, DE, MO) independently reviewed 50, randomly selected cardiopulmonary exercise tests. For each test, one (1) V-slope graph and one (1) Excess carbon dioxide graph were generated, identical to the data used to automatically determine ventilatory threshold. Investigators individually determined the ventilatory threshold using each of the methods. If an investigator believed the ventilatory threshold was indeterminate by either the V-Slope or Excess carbon dioxide method, they marked the ventilatory threshold for that method as “Not Determined”. The ventilatory threshold (mL/kg/min) values for the two methods were averaged for each individual investigator. Then the visually determined values for each investigator were compared to one another. If at least 2 investigators had ventilatory threshold values within 5% of one another, the ventilatory threshold values of the investigators were averaged and compared to the automated methods. If no 2 investigators had ventilatory threshold values within 5% of one another, the investigators reviewed the graphs together and determined the most representative value for ventilatory threshold. If investigators collectively felt ventilatory threshold was indeterminate for a given subject, the values determined through the automated method were reviewed and the most representative value of ventilatory threshold was determined by consensus. A paired-samples *t* tests were used to examine differences between the automated and visually determined ventilatory threshold methods. Two separate Intraclass Correlation Coefficients were used to examine the interrater reliability between investigators on visually determining ventilatory threshold and the relationship between the automated and visually determined ventilatory threshold methods.

*Results*

Investigators demonstrated excellent interrater reliability in visually determining ventilatory threshold (Intraclass Correlation Coefficients = .966 (95% Confidence Interval = 0.935-0.982); *p* < .001). The subjective analysis and the automated analysis Intraclass Correlation Coefficients also demonstrated excellent reliability (Intraclass Correlation Coefficients = .942 95% Confidence Interval = 0.493-0.982); *p* < .001).
